# Estimating the gestational age of spontaneous abortions identified via database algorithms: a literature review and empirical analysis in Norwegian register data

**DOI:** 10.1101/2025.03.20.25324313

**Authors:** Chaitra Srinivas, Jacqueline M. Cohen

## Abstract

**Introduction:** Spontaneous abortion is a common pregnancy outcome, but incomplete recording and missing gestational age in health databases pose challenges for research. Accurate timing of the start of pregnancy is critical information in drug safety studies.

**Objectives:** To review the literature on database algorithms to estimate gestational length for spontaneous abortions and clinical studies than can inform such algorithms. To estimate the average gestational age for algorithm-identified spontaneous abortions in Norway using interrupted time series analysis.

**Methods:** We used an algorithm to identify pregnancies registered in Norway from 2010-2020 and restricted to spontaneous abortions identified from registers of primary and specialist care, and births from the Medical Birth Registry of Norway. For births, we calculated the LMP by subtracting the recorded gestational age from the birth date. We assigned spontaneous abortions gestational ages ranging from 7 to 11 weeks and a corresponding LMP. We identified prescriptions from 70 days before to 97 days after LMP and calculated the number of antidepressant prescriptions per 10,000 pregnancies per day. We applied two-sample interrupted time series analysis with intervention points set at 28 and 55 days after LMP and compared antidepressant prescription trends after 28 gestational days for spontaneous abortions versus births.

**Results:** Database algorithms have used estimates for the gestational age at spontaneous abortion ranging from 8-10 weeks, and clinical studies suggest the mean or median gestational age at spontaneous abortion of around 9-10 weeks. In our interrupted time series analysis including 122,495 spontaneous abortions and 631,929 births, the 7-week assumption showed no post-intervention trend, suggesting underestimation. The 9-week assumption closely matched the trend for births (−0.051 prescriptions/day, 95% CI -0.090 to -0.013 vs. -0.056, 95% CI: -0.067 to - 0.046). The 8, 10, and 11-week assumptions showed less precise alignment. The best alignment occurred with the 64-day assumption (9.1 weeks).

**Conclusion:** Our study provides an empirically derived estimate for the average gestational age for algorithm-identified spontaneous abortions which can be applied in future research using the same pregnancy algorithm in Norway. While the 64-day estimate seems most accurate for our dataset, further validation studies are necessary to confirm its applicability in other contexts.

## Introduction

Miscarriage, or spontaneous abortion, is a common outcome of pregnancy.^1^ However, studying spontaneous abortions using secondary databases is challenging, as not all such pregnancies are recorded and the gestational length for these pregnancies is typically unavailable.^2,3^ Pregnancy may not be recognized prior to loss, especially in very early gestation, and only those cases for which medical attention is sought are recorded in the databases, which limits the comprehensiveness of the data. Despite this, many algorithms have been developed to identify pregnancies from sources such as insurance claims databases, electronic medical records, and national health registers.^4-8^ Furthermore, in connection with these algorithms a separate algorithm must be applied to estimate the gestational length and determine the start of pregnancy. In general, the algorithms are more accurate for term live births than for other pregnancy outcomes.^9-11^

Accurate gestational age classification is particularly important in pregnancy drug safety studies.^12-14^ Misclassification of gestational age in such studies can introduce differential exposure misclassification, affecting the validity of the findings.^15,16^ When gestational age is assumed rather than accurately recorded, it can reduce specificity at the individual level but may still approximate the population average. Accurate estimation of the average gestational age is essential to reliably capture medication use during pregnancy, particularly for medications that are frequently discontinued during pregnancy. If one overestimates the gestational length, then they risk counting pre-pregnancy days as part of the pregnancy (**Figure 1**). When misclassifying pre-pregnancy time as part of the exposure window, pre-pregnancy prescriptions may be considered as used during pregnancy. If the gestational length for spontaneous abortions is more likely to be misclassified, this could bias risk estimates positively, away from the null. If one underestimates the gestational length, we expect this could bias risk estimates toward the null, as one would be less likely to fill a prescription in the earliest weeks of pregnancy since they were, in fact, later in their pregnancy and may have already recognized they were pregnant. However, if we correctly time the start of pregnancy, on average, we should be able to validly estimate the risk.

**Figure 1.**
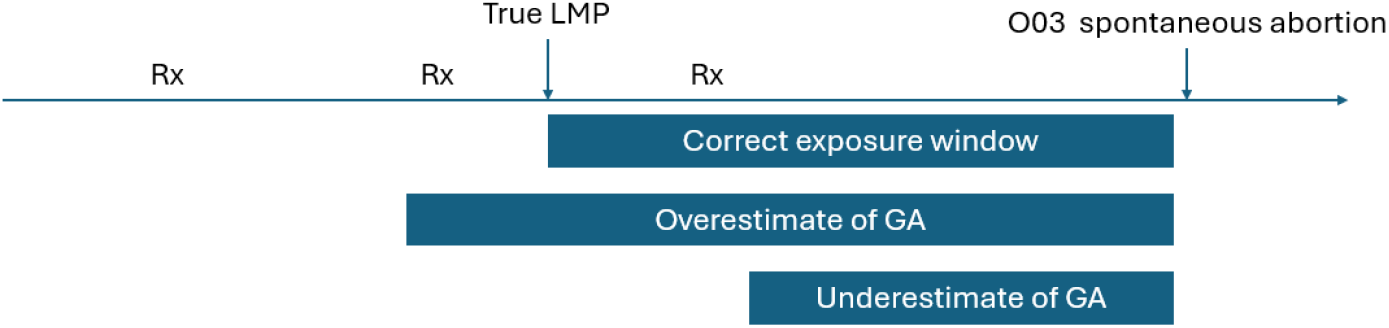
Systematic error in estimated gestational age (GA). If prescriptions are filled regularly until pregnancy is recognized, over and underestimation will affect capture of prescription fills. Rx denotes prescription fill and O03 is the ICD-10 code for spontaneous abortion. LMP, last menstrual period.

### Default gestational lengths assumed by existing databases algorithms

Previous studies have estimated gestational age for spontaneous abortions identified in healthcare databases using assumed mean or median values, rather than relying on clinical dating. These estimates are typically informed by prior clinical studies reporting the distribution of gestational ages at which spontaneous abortions occur. In 2006, Hornbrook et al. published an algorithm using multiple data sources from an integrated health system (Kaiser Permanente Northwest) in the US, incorporating diagnoses, procedures, pharmacy, and laboratory data, and validated the approach against medical records. For spontaneous abortions with no recorded gestational age or LMP, a national median gestational age of 10 weeks was applied as the default.^5^ Studies using US insurance claims data, such as those by Ailes et al., Korelitz et al., and MacDonald et al. identified pregnancy episodes using diagnostic and procedure codes, and assigned a gestational age informed by specific timing codes, or applied a default gestational age of 8 weeks for spontaneous abortions.^4,6,7^ For example, MacDonald et al. assigned a gestational age based on a specific timing code in 19% of spontaneous abortions and the rest were assigned the default. In 2024, Nordeng et al. published an algorithm for Norwegian register data and assigned gestational age using pregnancy markers where available, or 10 weeks for spontaneous abortions otherwise.^8^ They identified over 112,000 miscarriages and estimated a median gestational age of 10 weeks and 1 day as this algorithm still relied on default gestational age estimate for 66% of the miscarriages. Overall, most studies rely on an 8 to 10-week assumption for spontaneous abortions (**Table 1**), but vary in methods, settings, and levels of clinical validation. Given the lack of empirical validation of gestational age assumptions, there is a need for data-driven approaches to estimate gestational age.

**Table 1.** Assumed gestational age for spontaneous abortions identified in healthcare databases.

| Study | Gestational age |
| --- | --- |
| Hornbrook et al., 2006 | 10 weeks (median) |
| Ailes et al., 2016 | 8 weeks |
| Korelitz et al., 2013 | 8 weeks |
| MacDonald et al., 2019 | 8 weeks |
| Nordeng et al., 2024 | 10 weeks (median) |

### Clinical studies of the gestational age at spontaneous abortion

Some clinical studies have provided valuable insights into the gestational age at which spontaneous abortions occur, often relying on prospective cohort designs and self-reported data to estimate the timing. Wilcox et al. used data from the prospective “Menstruation and Reproduction History Study,” initiated in 1935.^17^ Freshman women at the University of Minnesota were invited to participate and instructed to record information related to their menstrual periods throughout their reproductive years. Between 1935 and 1939, 1,983 women volunteered, and 26 years later, a second cohort of women joined, raising the total to 3,889 participants. Spontaneous abortions were defined as reported pregnancies that terminated within 20 weeks of the previous menstrual period. After applying relevant exclusions, there were 4,346 pregnancies, of which 685 ended in spontaneous abortion. Cohorts 1 and 2 primarily differed in that women in cohort 1 had reached older ages. The mean gestational age of pregnancies ending in spontaneous abortion was 9.2 weeks and 9.4 weeks, respectively, in cohorts 1 and 2.

Mukherjee et al. used data from a prospective pregnancy cohort “Right from the Start” which recruited women who were pregnant or planning a pregnancy from different clinic- and community-based settings from 3 states in 2000-2009 to determine the risk of miscarriage among black and white women in US.^18^ Participants had an early pregnancy ultrasound and provided self-reported information on LMP. For most of the study, participants were required to enroll prior to 9 completed weeks. Pregnancy outcomes were self-reported and verified by medical records. Miscarriage was defined as a loss of a recognized pregnancy prior to 20 completed weeks of gestation. Miscarriage was documented in 537 (13%) of pregnancies during the study period and the median gestational age at the time of pregnancy loss for the cohort was 10 weeks.

Zhu et al. developed a hierarchical algorithm to estimate gestational age at the occurrence of spontaneous abortion or termination using US Medicaid data and validated the assigned gestational age using electronic medical records from Mass General Brigham hospital in Boston.^11^ The median gestational age of the 302 confirmed spontaneous abortions with valid gestational age at the end of pregnancy was 9.3 weeks.

### A pregnancy algorithm adapted to the Norwegian context

The Medical Birth Registry of Norway (MBRN) records virtually all births in Norway, plus spontaneous abortions from 12 gestational weeks.^19,20^ Additionally, induced abortions after 12 completed weeks require approval by a special medical board and are also registered in MBRN. However, spontaneous and induced abortions before 12 weeks are not registered, and in practice, pregnancies are not registered unless there is contact with a delivery hospital, which typically occurs at the time of the standard ultrasound at week 17-19.^21^ An algorithm was thus developed by Magnus et al. to utilize primary care data from the Norwegian Control and Payment of Health Reimbursements Database (KUHR) and specialist care data from the Norwegian Patient Registry (NPR) to identify additional pregnancies.^22-24^ The algorithm first identifies unique primary care (KUHR) registrations at least 90 days apart and specialist care (NPR) registrations at least 42 days apart. Then these are combined, prioritizing registrations from specialist care. Finally, these unique records are combined with records from the MBRN, removing invalid records of spontaneous or induced abortion from KUHR or NPR recorded during an ongoing pregnancy, or shortly following a registered pregnancy in the MBRN. Nordeng et al. have recently published a similar algorithm but required 56 days between unique pregnancy registrations.^8^ Neither gestational age nor LMP are recorded for spontaneous or induced abortions identified in the primary and specialist care data. Therefore, they developed an additional algorithm to assign gestational length based on the first time, on average, that various diagnosis codes occurred in pregnancies resulting in a birth recorded in the MBRN.

### Interrupted Time Series Analysis

It has previously been demonstrated that interrupted time series analysis can detect a decline in weekly prescription fills for antidepressants and ADHD medications as soon as a pregnancy may be recognized (4 weeks after LMP) which continues to decline until most pregnancies are recognized (7 weeks after LMP) among pregnancies resulting in a birth.^25^ In this study, we used interrupted time series analysis to test different gestational age assumptions. We examined antidepressant prescription fills to estimate the most realistic average gestational age at miscarriage, aiming to align the trends in prescription fills between 4 and 7 weeks after LMP for miscarriages with those for births.

### Aim

To carry out a series of two-sample interrupted time series analyses, comparing the trends in antidepressant prescription fills between 4 and 7 weeks after LMP for miscarriages identified from registers of primary and specialist care versus births from the medical birth register with recorded gestational age, using different assumptions about the mean gestational length for the miscarriages.

## Methods

We first identified all pregnancies from 2010-2020 using the pregnancy algorithm developed by Magnus et al. described above, with the modification that we did not include bleeding in pregnancy codes (ICD-10 O20.0; ICPC-2 W03). We excluded induced abortions, spontaneous abortions identified in 2020 for which we could not exclude the possibility that a birth was later registered, spontaneous abortions identified from the medical birth registry, and births from the medical birth registry with a missing gestational age.

For the births, we calculated the LMP date by subtracting the recorded best clinical estimate of gestational age in days from the date of birth. For the spontaneous abortions, we assigned an estimated gestational age ranging from 7 to 11 completed weeks.

We identified prescription fills for antidepressants (ATC codes starting with N06A) from the Norwegian Prescription Database (NorPD).^26^ We combined prescription fills for the same medication filled on the same day into one record. We calculated the number of antidepressant prescription fills per day from 70 days before LMP to 97 days after LMP. Then we scaled these to the number of prescriptions per day per 10,000 pregnancies using the relevant denominators (n=122,495 miscarriages; n=631,929 births).

We carried out two-sample interrupted time series analysis on the number of antidepressant prescription fills per day, where the “treatment” group were the spontaneous abortions, and the “controls” were the births. For each model, we set intervention points at days 28 and 55. We compared trends between spontaneous abortions and live births. We compared the linear post-intervention trends after 28 days and examined both the coefficients and 95% confidence intervals for the trends, and the differences between the trends, with the births serving as the reference group. We did not compare the linear trends after 55 days, as we did not expect to see similar trends in the group that was comprised of spontaneous abortions versus those that remained pregnant.

A Generalized Linear Model (GLM) was used with a Gaussian family and an identity link function, employing Newey-West standard errors, implemented in Stata SE 17.0.^27^

### Ethics/IRB Statement

The Regional Committee for Medical and Health Research Ethics of Norway (South-East A) gave ethical approval for this work (2017/2546)

## Results

The cohort included 913,816 pregnancies identified from KUHR, NPR, and MBRN 2010-2020, with 1% of spontaneous abortions and 2% of induced abortions identified from MBRN. We excluded 143,642 induced abortions, 1691 spontaneous abortions registered in MBRN, and 12,812 spontaneous abortions registered in KUHR and NPR in 2020. We further excluded 1376 (0.2%) births with no gestational age recorded.

When we examined the linear post-intervention trends for gestational age assumptions ranging from 7 to 11 weeks, we found the most similar trends for the 9-week assumption (Table 2, Figure 2). When we varied the number of days between 59 (8 weeks and 3 days) and 73 (10 weeks and 3 days), we found the most similar trend using the assumption of 64 days (9 weeks and 1 day).

**Table 2.** Comparison of linear postintervention trend in antidepressant prescriptions per day from 28 days after LMP with different mean gestational age assumptions for spontaneous abortions. Bold indicates the most similar trend.

|  | Coef. (95% CI) | P | Difference (95% CI) | P |
| --- | --- | --- | --- | --- |
| Control | -0.056 (-0.067, -0.046) | <0.001 | Reference | - |
| Spontaneous abortions (assumed mean gestational age in completed weeks) |  |  |  |  |
| 7 weeks | 0.183 (-0.029, 0.066) | 0.446 | 0.075 (0.026, 0.123) | 0.003 |
| 8 weeks | -0.037 (-0.075, 0.001) | 0.056 | 0.019 (-0.021, 0.059) | 0.347 |
| **9 weeks** | **-0.051 (-0.090, -0.013)** | **0.009** | **0.005 (-0.035, 0.045)** | **0.813** |
| 10 weeks | -0.047 (-0.088, -0.005) | 0.027 | 0.010 (-0.033, 0.052) | 0.663 |
| 11 weeks | -0.022 (-0.056, 0.012) | 0.197 | 0.034 (-0.002, 0.069) | 0.061 |
| Spontaneous abortions (assumed mean gestational age in days) |  |  |  |  |
| 59 days | -0.023 (-0.056, 0.010) | 0.179 | 0.034 (-0.001, 0.068) | 0.057 |
| 60 days | -0.023 (-0.054, 0.007) | 0.135 | 0.033 (0.001, 0.065) | 0.045 |
| 61 days | -0.023 (-0.054, 0.009) | 0.160 | 0.033 (0.000, 0.067) | 0.050 |
| 62 days | -0.033 (-0.058, -0.008) | 0.011 | 0.023 (-0.004, 0.050) | 0.092 |
| 63 days | -0.051 (-0.090, -0.013) | 0.009 | 0.005 (-0.035, 0.045) | 0.813 |
| **64 days** | **-0.053 (-0.089, -0.017)** | **0.004** | **0.004 (-0.034, 0.041)** | **0.854** |
| 65 days | -0.047 (-0.082, -0.011) | 0.010 | 0.010 (-0.028, 0.047) | 0.614 |
| 66 days | -0.038 (-0.078, 0.002) | 0.064 | 0.018 (-0.023, 0.060) | 0.387 |
| 67 days | -0.036 (-0.071, -0.001) | 0.046 | 0.020 (-0.017, 0.057) | 0.284 |
| 68 days | -0.023 (-0.059, 0.013) | 0.203 | 0.033 (-0.005, 0.070) | 0.086 |
| 69 days | -0.036 (-0.075, 0.004) | 0.078 | 0.021 (-0.020, 0.062) | 0.326 |
| 70 days | -0.047 (-0.088, -0.005) | 0.027 | 0.010 (-0.033, 0.052) | 0.663 |
| 71 days | -0.034 (-0.074, 0.006) | 0.093 | 0.022 (-0.019, 0.063) | 0.298 |
| 72 days | -0.028 (-0.067, 0.010) | 0.156 | 0.028 (-0.012, 0.068) | 0.176 |
| 73 days | -0.027 (-0.065, 0.011) | 0.161 | 0.029 (-0.010, 0.068) | 0.142 |

**Figure 2.**
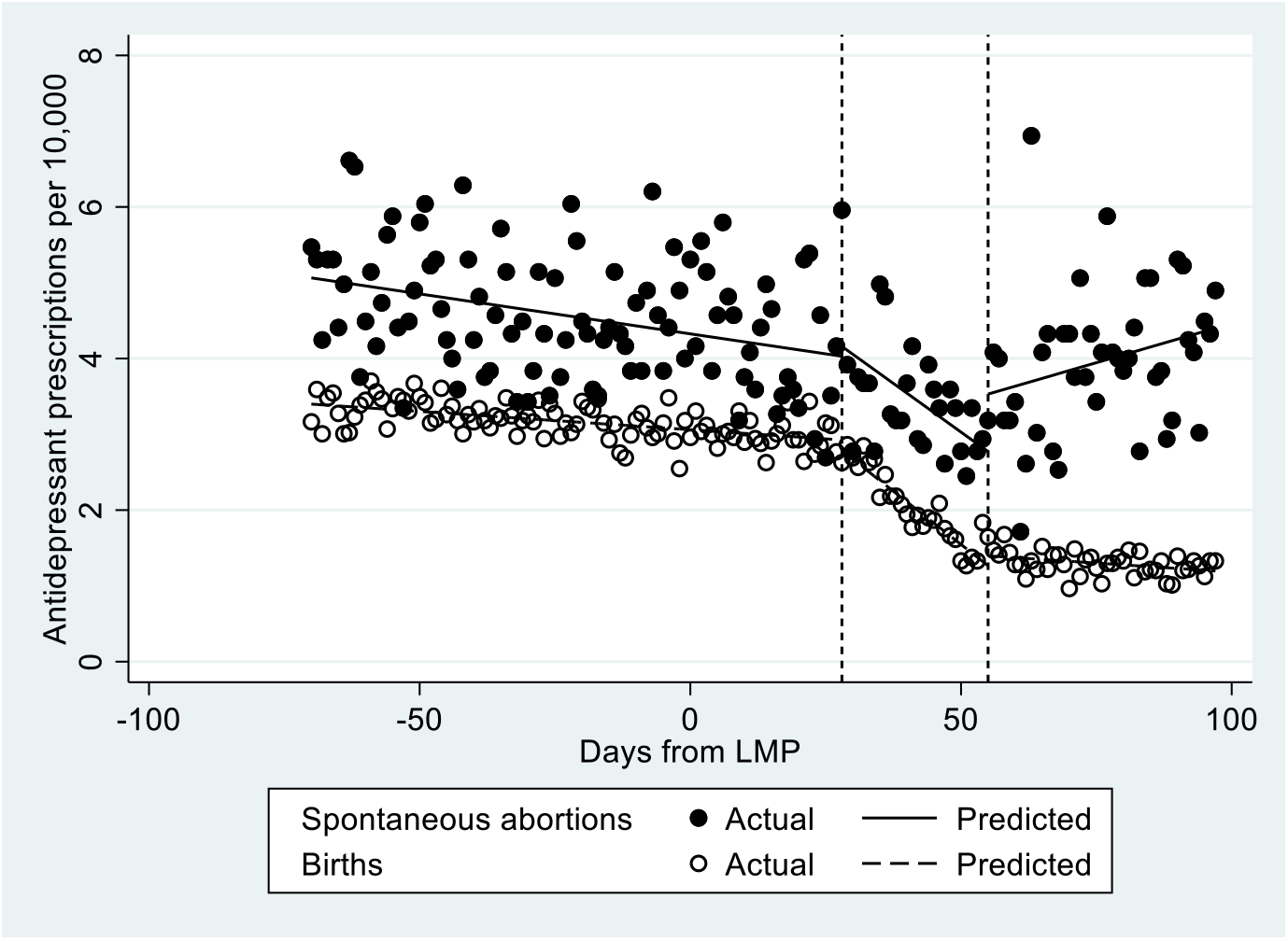
Number of antidepressant prescriptions per day per 10,000 pregnancies assuming a gestational length of 9 weeks for the spontaneous abortions. LMP, last menstrual period. Vertical dashed lines denote the intervention points specified in the model at 28 and 55 days after LMP.

## Discussion

Accurate estimation of gestational age is important for drug safety studies investigating risk of spontaneous abortion. Misclassifying gestational age can introduce bias and affect the validity of results. We found that previous algorithms for estimating gestational age for spontaneous abortions used estimates ranging from 8 to 10 weeks,^4-8^ and this is in line with clinical cohorts.^11,17,18^ In our study, we empirically assessed the most appropriate average estimate by comparing antidepressant prescription-fill trends in pregnancies, ultimately determining that 9.1 weeks (64 days) was our best estimate. This estimate can be applied to other studies in Norway and may offer a useful reference for gestational age assumptions in other settings.

Other timing algorithms for estimating gestational age are more detailed and could offer more precision in specific settings. However, whether the complexity of these algorithms offers significant benefits, is still unknown. Zhu et al. developed and validated a gestational age algorithm for spontaneous abortions using a Medicaid-insured pregnancy cohort linked to electronic medical records.^11^ Using a random forest approach with selected variables, the algorithm’s performance for spontaneous abortions was moderately accurate, with 58% of cases classified within 2 weeks of the gold standard (linked electronic medical records). In comparison, with a simpler approach with assignment of population median, they observed only a slightly lower accuracy (56%). This suggests that while more detailed algorithms may provide slightly improved estimates, their superiority remains uncertain. Our study’s focus on using a straightforward assumption aligns with a balance between simplicity and accuracy.

### Strengths

One of the key strengths of our study is the use of comprehensive data from both primary and secondary care, with 77% of spontaneous abortions registered in specialist care, an additional 22% registered in primary care, and just 1% registered in the medical birth registry. In Norway, cost is not a barrier to prenatal care, as healthcare is universally accessible and prenatal care is free, which further enhances the completeness and reliability of the data. Additionally, the high-quality prescription data covers all prescription fills for outpatients throughout the country and allows for precise tracking of antidepressant use during pregnancy. This strengthens the validity of our findings, as it minimizes biases related to incomplete prescription data.

### Limitations

Despite its strengths, our study has its limitations. First, the best estimate of gestational age that we derived may be specific to the Norwegian context. Different healthcare systems or algorithms might yield different results. We included spontaneous abortions identified in secondary care as early as 42 days after a prior pregnancy, but only up to 90 days after for pregnancies registered in primary care to enhance validity. This differs from the algorithm published by Nordeng et al. that allowed a shorter interval between pregnancy registrations from primary care,^8^ suggesting that the algorithms may capture slightly different pregnancy cohorts. However, our estimate of the mean gestational age of 9.1 weeks for spontaneous abortions was closely aligned with the mean gestational age reported in clinical cohorts from the US.^11,17^ Additionally, it may have been of interest to consider other medications that are frequently discontinued in pregnancy, such as ADHD medication, to compare the trends. However, while antidepressant use is relatively uncommon in Norway, other medications are used even less frequently. Therefore, we had limited power to assess other prescription-fill trends. Finally, we were unable to validate our assumption of the most accurate mean gestational age against medical records.

## Conclusions

Our study provides an empirically derived estimate of the average gestational age at miscarriage, which can be applied in future research using the same pregnancy algorithm in Norway. While the 64-day estimate is the most accurate for our dataset, further validation studies are necessary to confirm its applicability in other contexts.

## Data Availability

The data used in this study cannot be shared.

